# Attenuated GlycoRNA-microglial transcriptional coupling in the dorsolateral prefrontal cortex of patients with major depressive disorder: an exploratory interaction-based in-silico analysis

**DOI:** 10.64898/2026.10.12.26365369

**Authors:** Rui Xu, Yingqian Zhang, Michael Maes

## Abstract

**Background:** Glycosylated RNAs (GlycoRNAs) participate in immune recognition and intercellular communication^1^, but their relationship with brain cell-type transcriptional signatures in major depressive disorder (MDD) remains unclear. We examined GlycoRNA-related transcriptional coupling with microglial and other cellular signatures in the dorsolateral prefrontal cortex (DLPFC).

**Method:** Postmortem DLPFC transcriptomic data from 22 controls and 26 MDD patients (GSE102556)^2^ were analyzed. GlycoRNA-related pathway enrichment was quantified using gene set variation analysis (GSVA)^3^, whereas microglial, neuronal, astrocytic, and oligodendrocytic signatures were derived using single-sample gene set enrichment analysis (ssGSEA)^4–6^. Pearson correlations, linear regression, and generalized estimating equations (GEE) evaluated cell-type-specific associations and diagnostic interactions. Exploratory GEE results underwent false discovery rate (FDR) correction.

**Results:** No differentially expressed genes survived FDR correction, and GlycoRNA-related and cell-type signatures did not differ significantly between groups. In controls, GlycoRNA enrichment was positively associated with the microglial signature (β = 0.447, FDR-P = 0.006), with significantly weaker associations for neurons, astrocytes, and oligodendrocytes (all FDR-P < 0.05). The two-way GlycoRNA × is_microglia interaction was nominally significant (β = 0.210, P = 0.048), confirming that GlycoRNA transcriptional coupling is substantially stronger for microglia than for the other three cell types combined. The microglial association was numerically attenuated in MDD (β difference = −0.273, P = 0.167), but diagnostic interactions were nonsignificant.

**Conclusions:** GlycoRNA pathway transcriptional enrichment exhibits preferential microglial association in control DLPFC. It’s possible attenuation in MDD remains.

## 1. Introduction

Major depressive disorder (MDD) represents a highly prevalent and disabling psychiatric disorder worldwide, with substantial socioeconomic burden^7^. Accumulating human neuroimaging and post-mortem transcriptomic evidence consistently identifies dysfunction of the dorsolateral prefrontal cortex (DLPFC) as a core pathological feature of MDD, contributing to impaired emotion regulation and cognitive control^8–10^. Historically, mechanistic investigations of MDD have predominantly focused on neuronal apoptosis, monoamine signaling disturbance, and neuroinflammatory cascades^11,12^. Despite these advances, the biological underpinnings of MDD remain incompletely understood, and novel molecular layers of brain homeostasis await systematic exploration.

Glycosylated RNAs (GlycoRNAs), a newly discovered class of cell-surface-displayed RNA molecules covalently modified with N-linked glycans, participate in immune recognition and intercellular communication^1^. While GlycoRNAs have been characterized predominantly in immune cells and solid tissues, their transcriptional relationship with neuroimmune cell populations in the brain, particularly in the context of psychiatric disease, remains unexplored. The homeostasis of the brain microenvironment is coordinately maintained by four major cellular populations: neurons, astrocytes, oligodendrocytes, and microglia^13^. Among these cell types, microglia function as the primary resident immune cells of the brain and serve as critical orchestrators of neuroinflammation and neural circuit remodeling^14^.

Previous post-mortem brain transcriptomic studies of MDD have largely focused on independent group-wise differences in isolated signaling pathways or alterations in cellular composition/abundance. Few studies have characterized the rewiring of transcriptional correlations between molecular pathways and cellular activity states. Here, we hypothesized that the transcriptional relationship between GlycoRNA pathway processes and microglial signatures may be altered in the DLPFC of patients with MDD. We therefore examined group-specific associations between the GlycoRNA pathway transcriptional signature and transcriptional signatures of microglia, neurons, astrocytes, and oligodendrocytes.

To test this hypothesis, we performed a secondary bioinformatic reanalysis of publicly available post-mortem DLPFC transcriptomic data (GSE102556). Gene set variation analysis (GSVA) was applied to quantify the GlycoRNA pathway enrichment score, while single-sample gene set enrichment analysis (ssGSEA) was used to derive transcriptional activity scores for neurons, microglia, astrocytes, and oligodendrocytes. We further explored group-specific correlation patterns between GlycoRNA pathway transcriptional signature and cellular transcriptional signatures. This study tests whether the transcriptional coupling between GlycoRNA pathway processes and microglial signatures is altered in MDD DLPFC, offering a novel framework for interrogating neuroimmune dysregulation beyond conventional group-level differential expression.

## 2. Materials and Methods

### 2.1 Dataset acquisition

Public post mortem human DLPFC (Brodmann area 8/9) transcriptomic dataset GSE102556 was retrieved from the NCBI GEO repository^15^. This dataset was originally generated by Labonté et al.^2^. Briefly, post mortem brain specimens were collected from human autopsy cohorts, with clinical diagnoses established using medical records and psychological autopsy information following DSM IV criteria. Subjects with known neurodegenerative disease, heavy alcohol or substance use disorder were excluded in the original study. Clinical metadata including age, sex, post mortem interval, brain tissue pH, RNA integrity number, antemortem medication exposure and cause of death were available for each individual sample. Demographic and clinical characteristics of the included samples are summarized in Supplementary Table 1. The original cohort contained 48 subjects with multi region brain RNA seq data. For our present secondary analysis, only DLPFC samples were retained. After sample metadata matching and quality filtering, 48 DLPFC samples consisting of 22 non psychiatric sudden death controls and 26 individuals diagnosed with major depressive disorder (a subset of MDD cases died by suicide) were included for downstream bioinformatic re analysis. No raw sequencing data were generated in this study.

### 2.2 Data preprocessing

FPKM expression matrix was loaded and processed in R (v4.3.2). Genes with consistently low expression were filtered out, retaining genes detected with FPKM >2 in at least 10 samples. Expression values were log_2_ transformed (log_2_(FPKM+1)) for subsequent analyses to avoid infinite values from zero counts. Ensemble gene identifiers were converted to official HUGO gene symbols using ‘org.Hs.eg.db’ and ‘AnnotationDbi Bioconductor’ packages. Only genes mapped to valid gene symbols were kept for further analysis. Sample metadata were matched between expression matrix and phenotype table to ensure sample consistency.

### 2.3 Differential expression analysis

Differential expression between MDD and control groups was performed using the ‘limmà R package with empirical Bayes moderation^16^. Model matrix was constructed with group status as the primary explanatory variable. Contrast was defined as MDD versus control. Genes satisfying thresholds of nominal P value < 0.05 and |log_2_ fold change| > 0.25 were considered nominally differentially expressed genes (DEGs). False discovery rate (FDR) was applied for multiple testing correction. Of note, no genes reached statistical significance after FDR correction.

### 2.4 GlycoRNA pathway transcriptional signature evaluation

A curated GlycoRNA glycosylation related gene set was assembled according to previously reported human GlycoRNA associated glycosyltransferase genes^1^. The ‘gsva()’ function from the GSVA R package was applied with ‘method = ‘gsva’’ to compute sample wise GlycoRNA pathway enrichment scores, which served as a proxy for GlycoRNA pathway transcriptional signature^3^.

### 2.5 Cell-type signature quantification

Canonical cell type marker gene sets for microglia, neurons, astrocytes, and oligodendrocytes were curated from well established human and rodent CNS cell specific transcriptomic resources^4^. Single sample gene set enrichment analysis (ssGSEA, ‘method=‘ssgsea’’ within GSVA package) was implemented to calculate cell type enrichment scores for each individual sample, representing inferred transcriptional signature of the four major central nervous system cell populations^6^. Genes absent from the expression matrix were excluded from each gene set before ssGSEA computation.

### 2.6 Statistical analysis

All statistical analyses were performed in R v4.3.2. The Wilcoxon rank sum test compared GSVA/ssGSEA enrichment scores between MDD and control groups. Pearson correlation assessed pairwise associations between the GlycoRNA pathway score and each cell type signature across all samples, controls, and MDD patients. Separate ordinary least squares regression models with a two way interaction term (‘GlycoRNA_score × group’) were fitted per cell type to test whether diagnosis moderated these correlations.

To evaluate the three way interaction GlycoRNA score × group × cell type, data were converted to long format (four rows per subject, one for each cell type ssGSEA score) and analysed via generalized estimating equations (GEE; ‘geepack’ package)^17^. We specified a Gaussian family, independent working correlation structure, and subject level clustering to handle repeated measures, with microglia and control group set as reference levels. Post hoc contrasts calculated the MDD versus CTRL difference in GlycoRNA cell signature slope within each cell type, with FDR correction for multiple testing. The delta method generated 95 % confidence intervals for predicted simple slopes. Sensitivity GEE analyses used square root and log transformed cell type scores. Complementary dummy variable GEE models contrasted microglia against the pooled other three cell types, testing two way and three way interaction terms.

Nominal significance was defined as P < 0.05; P values 0.05–0.10 were considered marginally significant.

### 2.7 Visualization

Volcano plot for differential genes, boxplots for GSVA scores, scatter plots for correlation relationships, and correlation coefficient heatmap were generated using ‘ggplot2’, ‘ggpubr’ and ‘pheatmap’ packages. For GEE outputs, forest plot was plotted to visualize regression coefficients and 95 % confidence intervals of key interaction terms. Faceted simple slope plot with delta method derived 95 % confidence bands was generated to visualize predicted relationships across group and cell type strata. All graphical outputs were exported as vector compatible PDF files.

## 3. Results

### 3.1 Differential gene expression in MDD prefrontal cortex

After filtering and quality control, 13,826 expressed genes were retained across 48 DLPFC samples (22 non-psychiatric controls, 26 MDD cases). Applying nominal-level thresholds (nominal P < 0.05 and |log_2_ fold-change| > 0.25), limma differential expression analysis identified 122 up-regulated and 46 down-regulated genes in MDD relative to controls. However, no gene survived FDR correction (minimum adjusted P = 0.777), indicating that the magnitude of transcriptomic perturbation in this post-mortem cohort was modest and no single gene reached genome-wide statistical significance. The top nominally altered genes included several non-coding RNAs and protein-coding genes with small effect sizes (|log_2_FC| range: 0.25-0.47). Full differential expression results are provided in Supplementary Table 2, and the transcriptome-wide distribution is visualized in the volcano plot (Figure 1).

**Figure 1.**
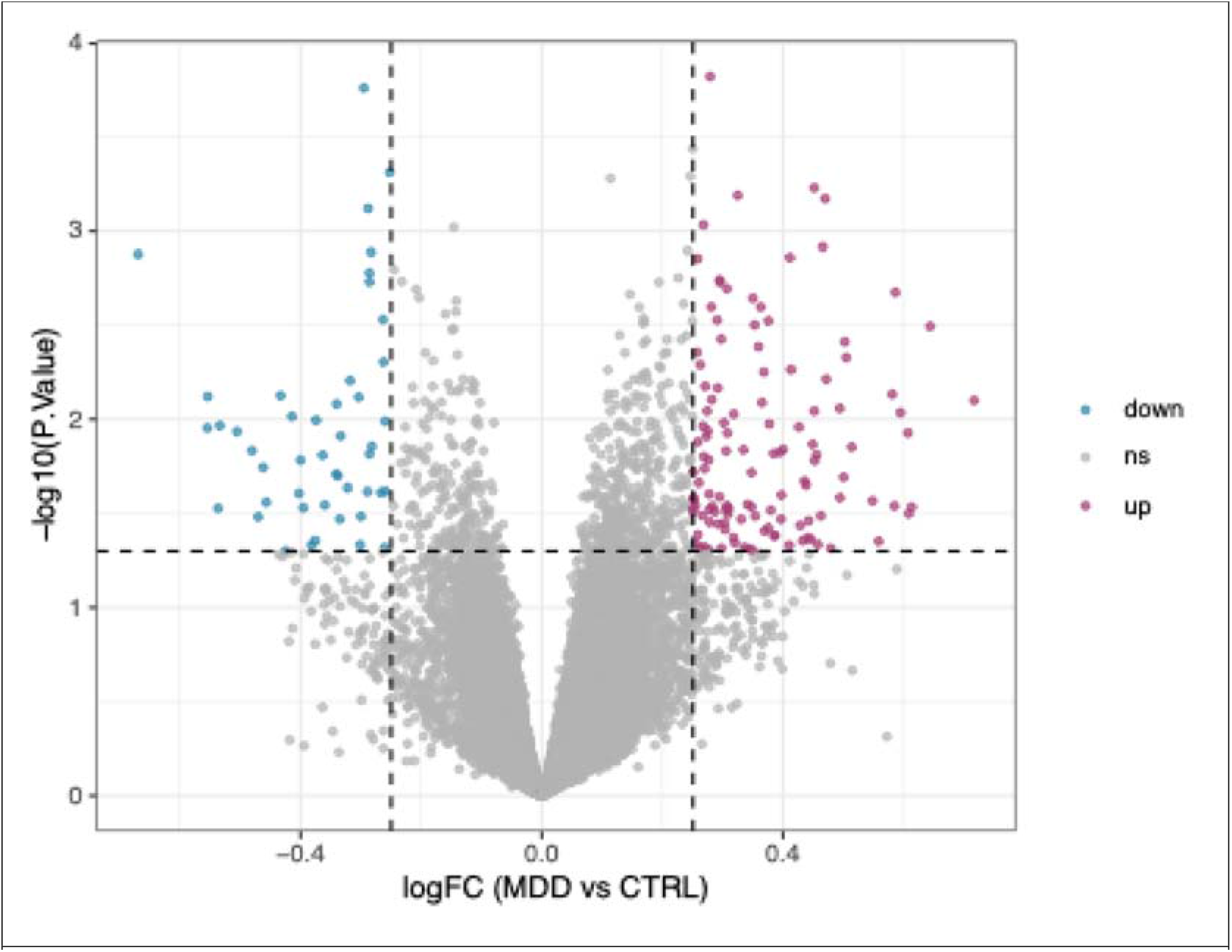
Differential gene expression in the prefrontal cortex of MDD patients versus controls

### 3.2 GlycoRNA pathway and cell-type signatures show no group-level differences

Global GlycoRNA pathway transcriptional signature, quantified by GSVA enrichment score, did not differ significantly between MDD patients and controls (mean ± SD: CTRL = 0.025 ± 0.155, MDD = −0.023 ± 0.165; Wilcoxon rank-sum test, P = 0.664) (Supplementary Table 3). Similarly, ssGSEA-derived cell-type signatures for microglia (P = 0.725), neurons (P = 0.741), astrocytes (P = 0.432), and oligodendrocytes (P = 0.185) showed no significant between-group differences. These results indicate that MDD pathology in this DLPFC cohort is not accompanied by global shifts in GlycoRNA pathway signature or macroscopic cell-type abundance signatures (Figure 2; Supplementary Figures 1). Per-sample scores are provided in Supplementary Table 4.

**Figure 2.**
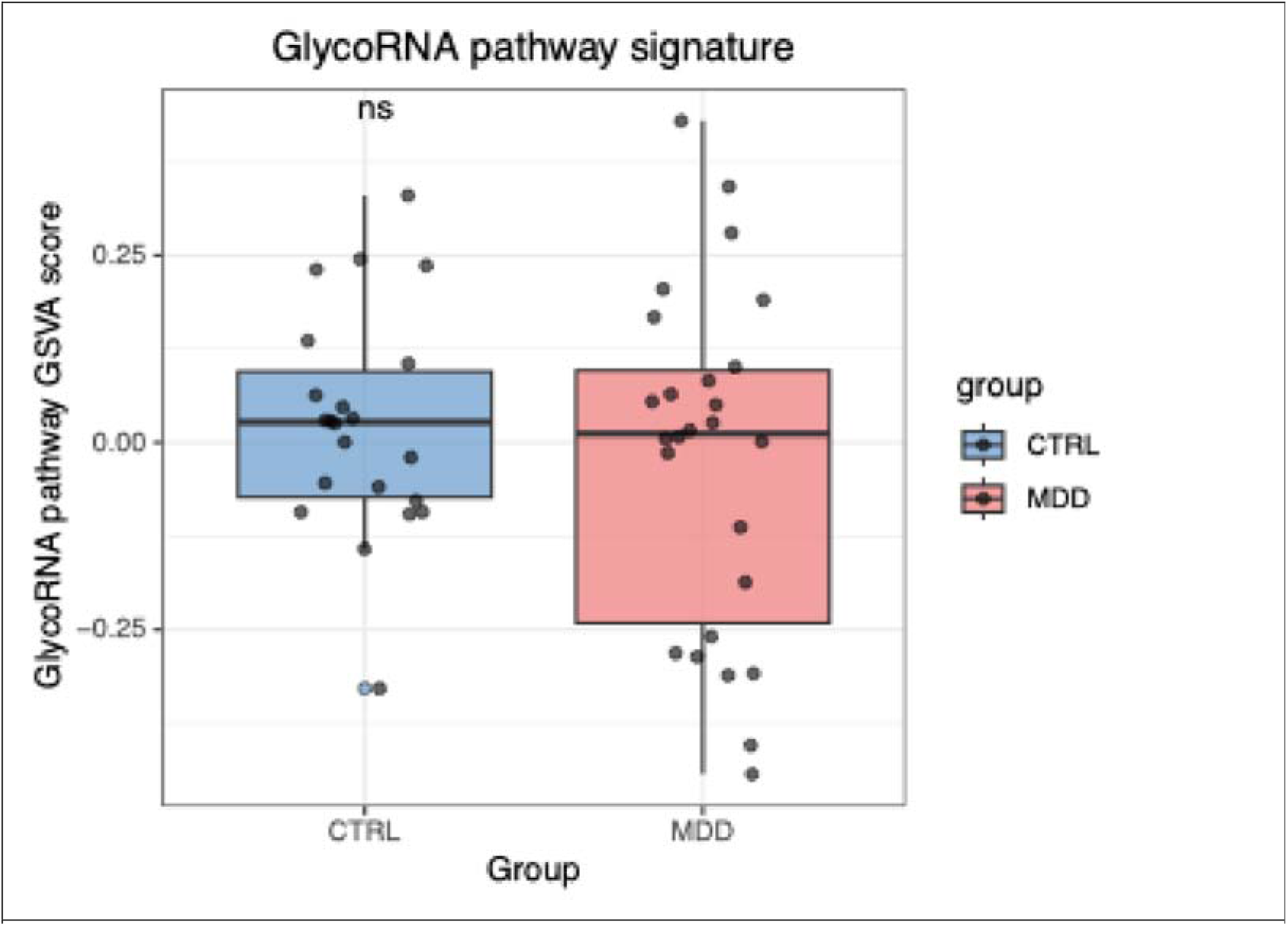
GlycoRNA pathway GSVA signature in MDD versus control groups.

### 3.3 GlycoRNA signature is positively correlated with microglial, but not neuronal, astrocytic, or oligodendrocytic, signatures

Pearson correlation analysis revealed a marginally positive association between GlycoRNA pathway score and microglial ssGSEA score across all samples (r = 0.282, P = 0.052). Stratified subgroup analysis showed that this association was numerically stronger in controls (r = 0.396, P = 0.068) and attenuated in MDD patients (r = 0.251, P = 0.216), although neither subgroup correlation reached formal significance (Figure 3).

**Figure 3.**
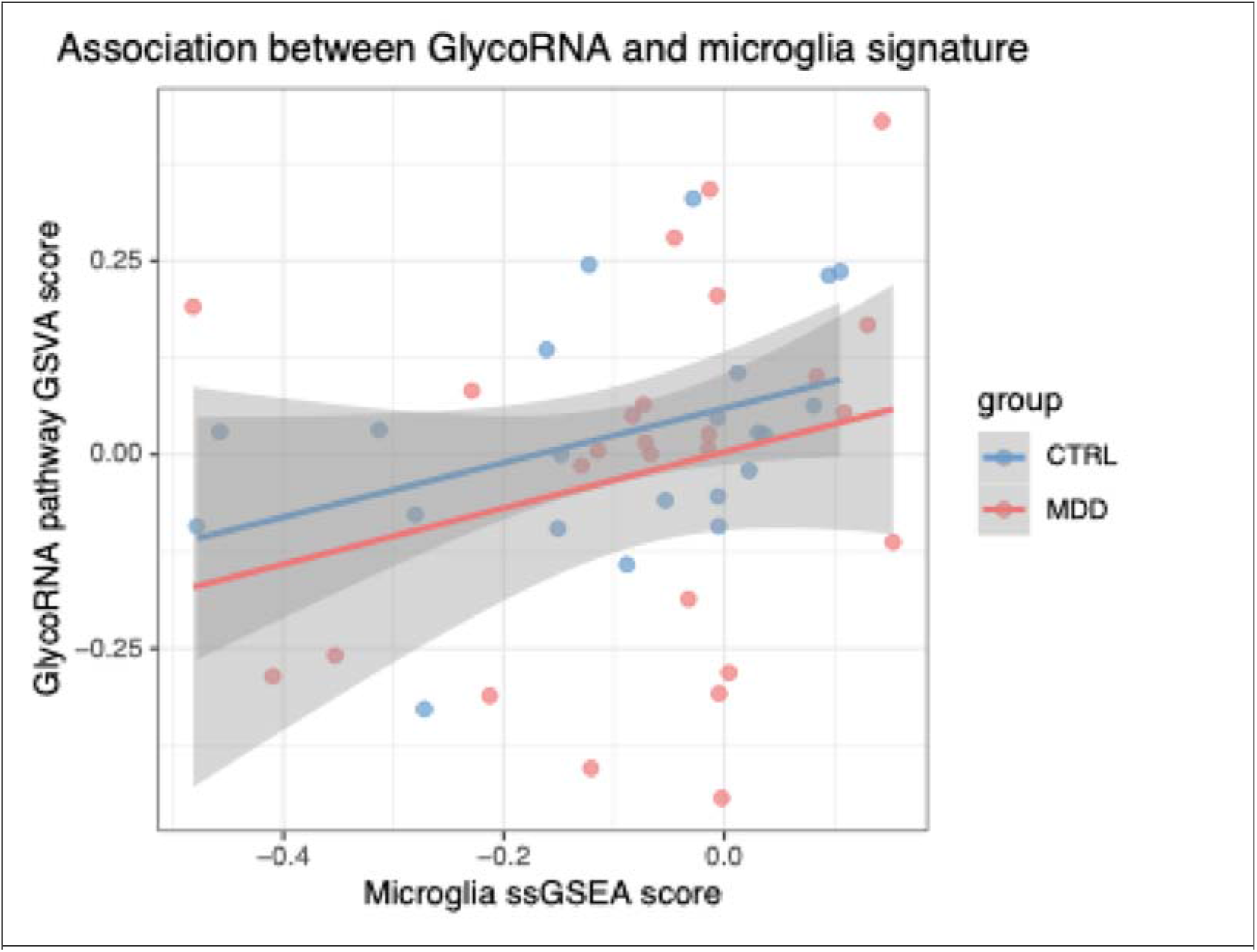
Association between GlycoRNA pathway GSVA score and microglia ssGSEA enrichment score

No significant correlations were observed between GlycoRNA score and neuronal (all samples: r = 0.124, P = 0.401; CTRL: r = 0.003, P = 0.988; MDD: r = 0.203, P = 0.320), astrocytic (all: r = 0.116, P = 0.434; CTRL: r = −0.006, P = 0.980; MDD: r = 0.178, P = 0.384), or oligodendrocytic signatures (all: r = 0.208, P = 0.157; CTRL: r = 0.034, P = 0.880; MDD: r = 0.270, P = 0.182) (Supplementary Figures 2, Supplementary Table 5). Among the four cell types examined, microglia showed the strongest GlycoRNA-cell-type coupling, particularly in control subjects. Correlation coefficients and P-values across all strata are summarized in the heatmap (Figure 4).

**Figure 4.**
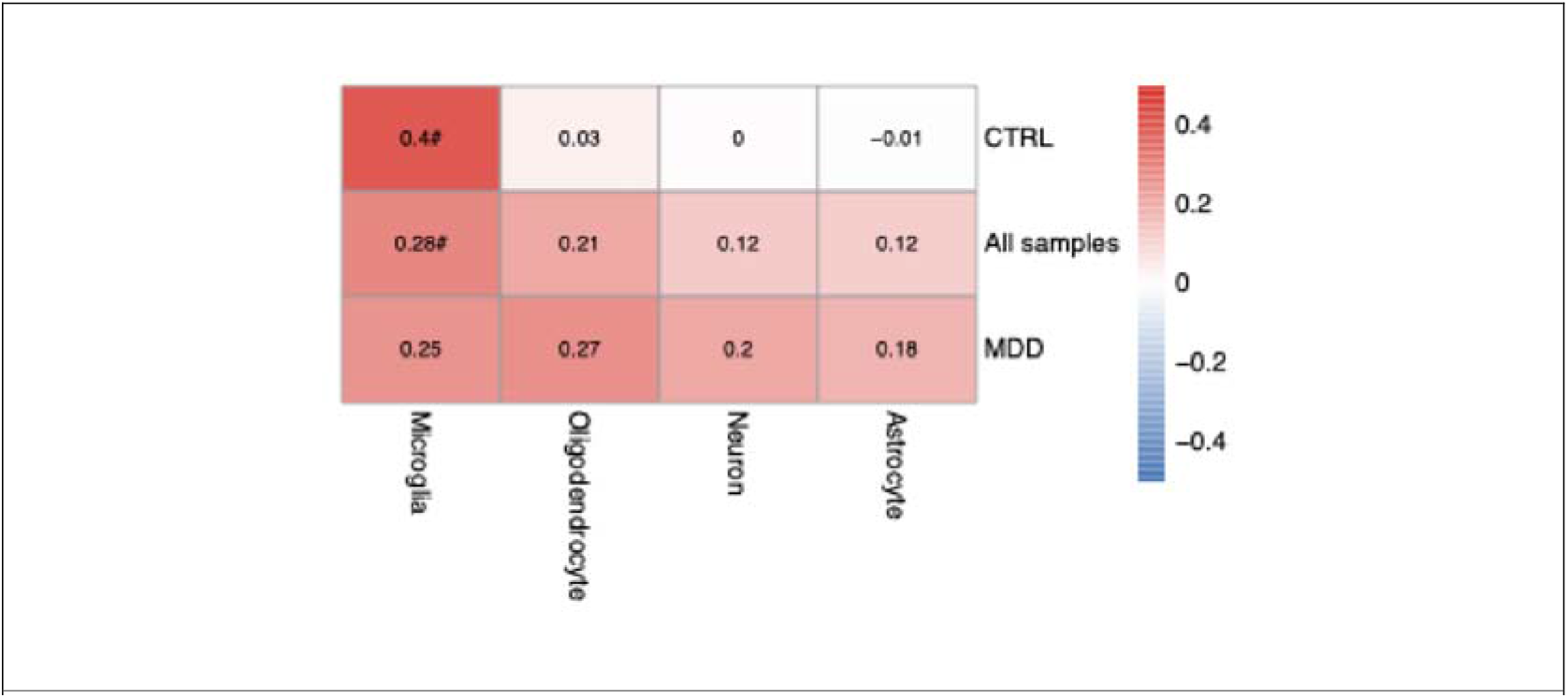
Pearson correlation coefficients between GlycoRNA pathway GSVA score and ssGSEA scores of four brain cell types. “*” denotes nominally significant correlation (P < 0.05); “#” indicates marginally significant correlation (0.05 ≤ P < 0.1).

### 3.4 Two-way interaction linear regression per cell type

To formally test whether diagnosis moderates the GlycoRNA-cell-type relationship, separate ordinary least-squares regression models with a GlycoRNA × group interaction term were fitted for each cell type. None of the two-way interaction terms reached nominal significance: microglia (β= −0.273, P = 0.316), neurons (β = 0.007, P = 0.610), astrocytes (β = 0.016, P = 0.608), or oligodendrocytes (β = 0.053, P = 0.550). Thus, although the GlycoRNA-microglia slope was numerically lower in MDD than in controls, per-cell-type ordinary regression did not establish a statistically significant diagnostic difference (Supplementary Table 6).

### 3.5 GEE three-way interaction: ‘GlycoRNA × group × cell typè

To jointly model the GlycoRNA-cell-type slope across all four cell types while accounting for within-subject repeated measures, a primary GEE model (cell_score ∼ GlycoRNA × group × celltype) was fitted on long-format data (192 observations from 48 subjects × 4 cell types).

In the reference stratum (control subjects, microglia as reference cell type), GlycoRNA score showed a significant positive main effect on microglial signature (β = 0.447, Wald P = 0.002, FDR-adjusted P = 0.006). This indicates that in non-psychiatric control DLPFC, higher GlycoRNA pathway transcriptional signature is robustly associated with higher microglial signature.

Relative to the microglial slope in controls, the GlycoRNA slope was significantly weaker for neurons (β = −0.447, P = 0.003, FDR = 0.006), astrocytes (β = −0.448, P = 0.001, FDR = 0.004), and oligodendrocytes (β = −0.436, P = 0.007, FDR = 0.013). These cell-type-specific interaction terms confirm that GlycoRNA transcriptional coupling is preferentially associated with microglial rather than neuronal, astrocytic, or oligodendrocytic signatures in the healthy brain.

Critically, the two-way GlycoRNA × group term (which tests whether the microglial GlycoRNA slope differs between MDD and controls) was negative in direction and of notable magnitude (β = −0.273), indicating a ∼61% attenuation of the GlycoRNA-microglia slope in MDD relative to controls, but did not reach nominal significance (P = 0.167, FDR = 0.222).

All three-way interaction terms (GlycoRNA × groupMDD × celltype) were non-significant: neuron (P = 0.166), astrocyte (P = 0.122), and oligodendrocyte (P = 0.112). The full coefficient table is provided in Supplementary Table 7, and the forest plot of key terms is shown in Figure 5.

**Figure 5.**
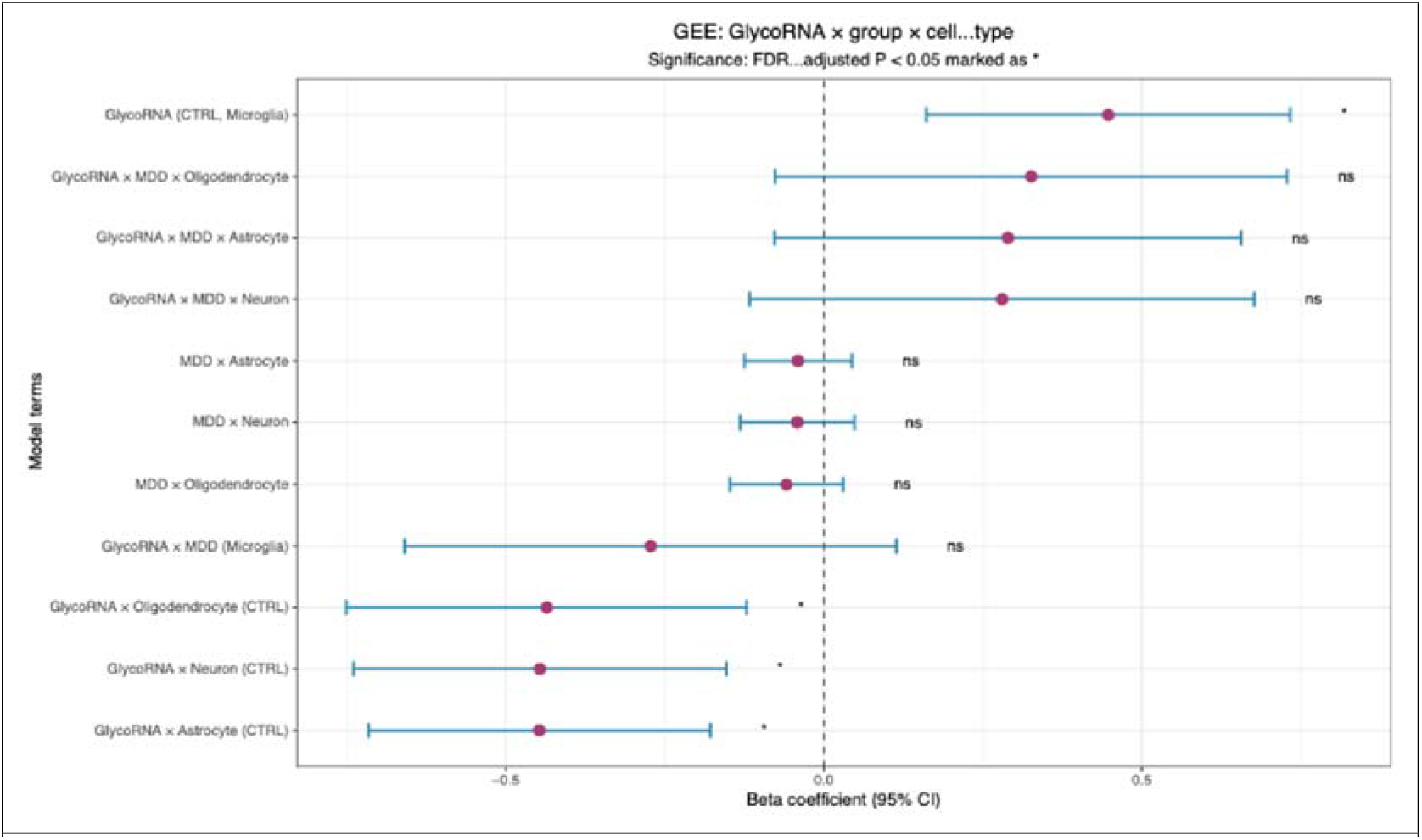
Forest plot of GEE three-way interaction model coefficients.

### 3.6 Post hoc slope contrasts within each cell type

Planned post-hoc contrasts directly estimated the MDD-minus-control difference in GlycoRNA slope within each cell type (Table 1). Consistent with the overall pattern, microglia showed the largest magnitude of slope reduction in MDD (Δ slope = −0.273, P = 0.167, FDR = 0.517), whereas neurons (Δ = 0.007, P = 0.468), astrocytes (Δ = 0.016, P = 0.517), and oligodendrocytes (Δ = 0.053, P = 0.387) showed near-zero or positive slope differences. None of the within-cell-type contrasts survived FDR correction. The faceted simple-slope plot with delta-method 95% confidence bands visually confirmed that only microglia displayed an apparent attenuation of positive GlycoRNA slope from CTRL toward MDD, while the other three cell types showed near-flat slopes in both groups (Figure 6).

**Table 1.** Post-hoc MDD-minus-control GlycoRNA slope contrast within each cell type.

| Cell type | $\Delta$ slope (MDD minus CTRL) | SE | Z | RAW P | FDR adjusted P |
| --- | --- | --- | --- | --- | --- |
| Microglia | $-0.273$ | 0.197 | $-1.383$ | 0.167 | 0.517 |
| Neuron | 0.007 | 0.010 | 0.725 | 0.468 | 0.517 |
| Astrocyte | 0.016 | 0.025 | 0.648 | 0.517 | 0.517 |
| Oligodendrocyte | 0.053 | 0.061 | 0.865 | 0.387 | 0.517 |

**Figure 6.**
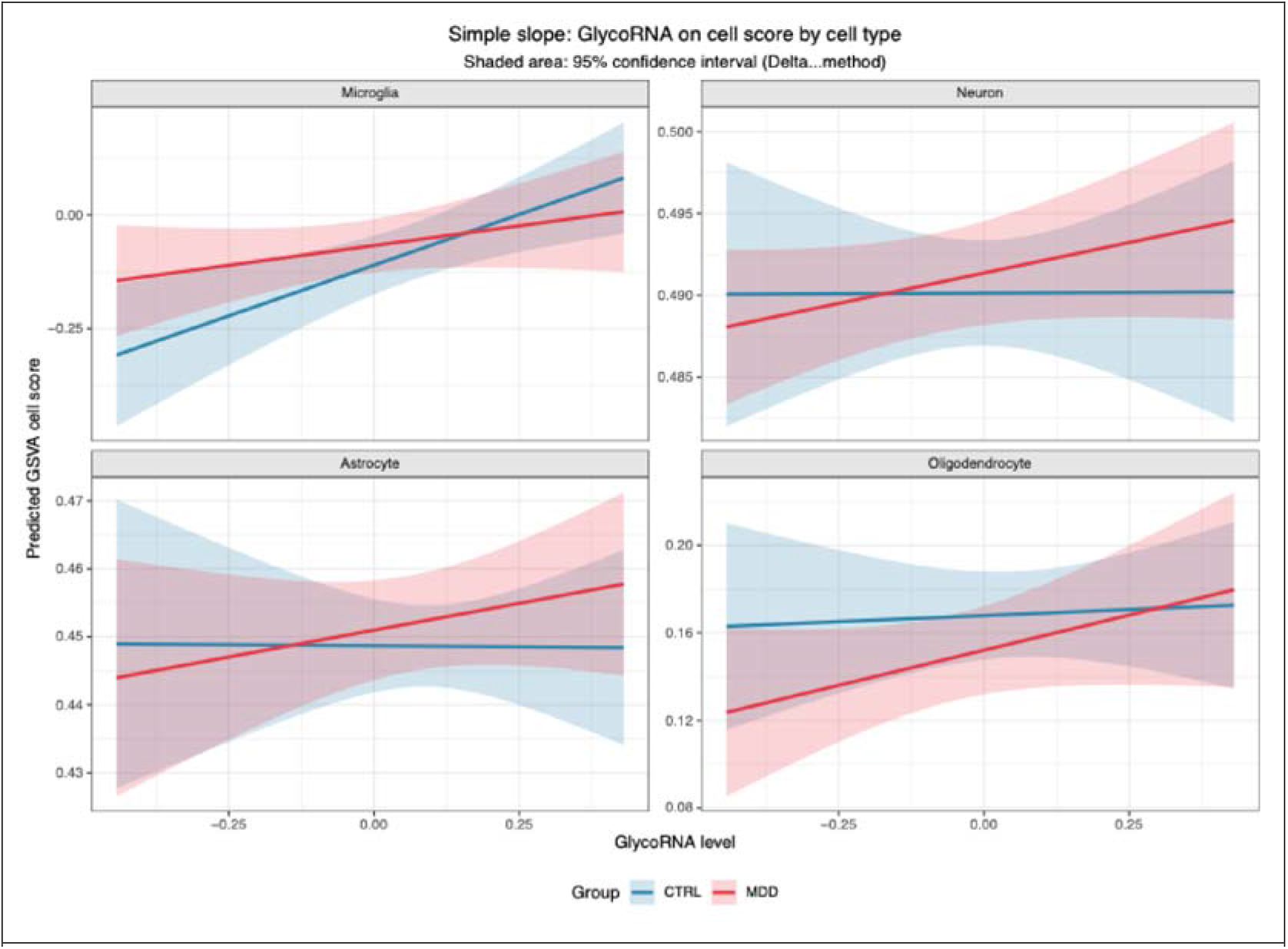
Predicted simple slopes of GlycoRNA pathway score on cell-type signature, stratified by diagnostic group and cell type.

### 3.7 Sensitivity analyses: transformed outcomes

To assess robustness against outcome distribution assumptions, identical three-way GEE models were refitted on square-root-transformed and log-transformed cell-type scores. Under both transformations, the GlycoRNA main effect on microglial signature remained significant (sqrt: β = 0.424, P = 0.007; log: β = 2.052, P = 0.065), and all three-way interaction terms remained non-significant (sqrt: neuron P = 0.146, astrocyte P = 0.127, oligodendrocyte P = 0.114; log: neuron P = 0.148, astrocyte P = 0.144, oligodendrocyte P = 0.136). The direction and relative magnitude of effects were consistent with the primary model, indicating that the observed pattern was not driven by outcome distributional properties (Supplementary Tables 8).

### 3.8 Dummy-variable GEE: microglia versus pooled other cell types

As a complementary microglia-focused test, we fitted GEE models using a binary is_microglia dummy (1 = microglia, 0 = pooled neurons + astrocytes + oligodendrocytes).

The two-way GlycoRNA × is_microglia interaction was nominally significant (β = 0.210, P = 0.048, FDR = 0.060), confirming that GlycoRNA transcriptional coupling is substantially stronger for microglia than for the other three cell types combined. The three-way GlycoRNA × group × is_microglia term, which specifically tests whether the diagnostic decoupling is microglia-selective, was directionally consistent (β = −0.298, P = 0.128, FDR = 0.255) but did not reach significance (Supplementary Tables 9).

## 4. Discussion

### 4.1 Principal findings

In this secondary analysis of post-mortem DLPFC transcriptomes from 22 non-psychiatric controls and 26 MDD individuals, we report three principal findings. First, no transcriptome-wide differentially expressed genes survived FDR correction, and neither GlycoRNA pathway transcriptional signature nor inferred microglial, neuronal, astrocytic, or oligodendrocytic signatures differed globally between MDD and control groups. Second, and most notably, GlycoRNA pathway transcriptional signature was robustly and specifically coupled to microglial transcriptional signature in the control brain, a relationship confirmed by a significant GEE main effect (β = 0.447, FDR-adjusted P = 0.006) and a nominally significant microglia-vs-other-cells dummy contrast (P = 0.048). Third, this GlycoRNA-microglial slope was numerically attenuated in MDD by approximately 61%, a directionally consistent pattern observed across ordinary regression, three-way GEE, transformed-outcome sensitivity analyses, and microglia-focused dummy models, although the formal three-way interaction did not reach statistical significance in this moderate-sized cohort. Collectively, these data support a hypothesis-generating model in which physiological GlycoRNA-microglial transcriptional coordination in the prefrontal cortex becomes decoupled in MDD.

### 4.2 GlycoRNA-microglial coupling in the healthy DLPFC: a neuroimmune glycosylation axis

The significant positive association between GlycoRNA pathway signature and microglial signature in control subjects (β = 0.447, FDR = 0.006) is biologically coherent^18^. GlycoRNAs are cell-surface-displayed RNAs covalently modified with glycosylation machinery, originally characterized in immune cells where they serve as functional ligands for selectins and Siglec receptors, and subsequently detected across mammalian tissues^1,19^. The glycosyltransferase gene set used here encodes the canonical ER/Golgi glycosylation machinery required for glycoRNA biosynthesis, comprising GALNT, B4GALT, ST6GAL, ST3GAL, and SLC35A family enzymes^1^. Microglia, as the resident innate immune cells of the CNS, are characteristically rich in surface glycans that mediate lectin recognition, complement receptor engagement, phagocytic synapse formation, and cytokine receptor glycosylation-dependent signaling^20,21^. It is therefore plausible that higher transcriptional signature of the GlycoRNA/glycosylation machinery in bulk DLPFC tissue tracks specifically with the transcriptional state of microglia rather than with neurons, astrocytes, or oligodendrocytes, whose cell-surface glycosylation profiles and glycosyltransferase repertoires are functionally distinct^22^.

The GEE model further demonstrated that the GlycoRNA slope was significantly weaker for neurons (β = −0.447, FDR = 0.006), astrocytes (β = −0.448, FDR = 0.004), and oligodendrocytes (β = −0.436, FDR = 0.013) relative to microglia. This cell-type specificity, rather than a general correlation with all CNS cell populations, strengthens the interpretation that the GlycoRNA signature reflects a microglia-associated glycosylation program rather than a nonspecific marker of tissue cellularity or overall transcriptional signature. The complementary dummy-variable model, which contrasted microglia against the pooled other three cell types, reached nominal significance for the GlycoRNA × is_microglia term (β = 0.210, P = 0.048, FDR = 0.060), independently corroborating this microglial selectivity.

### 4.3 Attenuated GlycoRNA-microglial coupling in MDD: a hypothesis of neuroimmune glycan decoupling

Although the GlycoRNA-microglial slope did not differ significantly between MDD and controls at the formal statistical level, the magnitude and direction of the effect are biologically noteworthy. The estimated slope reduction of −0.273 units corresponds to an approximately 61% attenuation of the positive GlycoRNA-microglial relationship in MDD relative to controls (0.447 → 0.174). This pattern was consistent across multiple analytic frameworks: (i) per-cell-type ordinary least-squares interaction regression showed the largest interaction effect for microglia (β = −0.273, P = 0.316); (ii) the GEE two-way GlycoRNA × group term for microglia was negative and of comparable magnitude (β = −0.273, P = 0.167); (iii) post-hoc within-cell-type slope contrasts identified microglia as the only cell type showing a negative MDD-minus-control slope difference; (iv) square-root- and log-transformed sensitivity models reproduced the same directional attenuation; and (v) the microglia-vs-other dummy three-way term was directionally consistent (β = −0.298, P = 0.128).

We interpret this convergent directional pattern as evidence that the physiological transcriptional coordination between GlycoRNA/glycosylation machinery and microglial identity may become disrupted in MDD DLPFC. Several mechanistic scenarios could account for such decoupling. First, MDD-associated microglial activation or dystrophy may alter the relationship between glycosyltransferase expression and microglial identity markers—for instance, primed or reactive microglia may upregulate certain glycosylation pathways independently of their baseline transcriptional identity^18,23^, or may downregulate surface glycan structures as a consequence of chronic inflammatory signaling^24^, a scenario consistent with the non-inflammatory but altered microglial phenotype observed in MDD post-mortem brain^25^. Second, medication exposure (antidepressants, antipsychotics) in MDD cases may directly modulate glycosylation enzyme expression or microglial activation state^26–28^, uncoupling the normally tight regulatory relationship between glycosylation machinery and microglial identity. Third, MDD neuropathology may involve altered glycan composition on microglial surface (e.g., hyposialylation, altered branching of N-glycans)^18,24^, a scenario directly supported by the recent finding that chronic stress reduces ST3GAL1-mediated sialylated O-glycosylation in the mPFC and that this glycan alteration is causally linked to depressive-like behavior^29^. Fourth, suicide-subtype specificity within the MDD cohort may introduce heterogeneity that dilutes the group-level signal.

Importantly, the absence of a statistically significant three-way interaction should be interpreted in the context of this cohort’s statistical power. With only 48 subjects and 192 long-format observations, the GEE model, while appropriate for jointly modeling repeated cell-type measures, has limited power to detect higher-order three-way interactions, which typically require substantially larger samples. The observed effect size (Δ slope ≈ −0.273, corresponding to a ∼61% slope reduction) is, however, biologically meaningful, and its consistency across analytic strata argues against it being a chance finding. We therefore frame this as an exploratory, hypothesis-generating result rather than a definitive conclusion.

### 4.4 Why no global transcriptomic or pathway-level differences?

A notable feature of our results is the near-absence of between-group differences at the level of individual genes (zero genes surviving FDR correction) and the absence of group-level differences in GlycoRNA pathway score or any cell-type signature. This is consistent with prior post-mortem MDD transcriptomic studies, which have generally reported modest, often FDR-non-significant, gene-level changes in prefrontal cortex, particularly when comparing mixed MDD cohorts against non-psychiatric sudden-death controls^30,31^. Several factors likely contribute. First, bulk RNA-seq from post-mortem tissue masks cell-type-specific changes; a molecular alteration restricted to microglia (which constitute only ∼5% of cortical cells)^32^ may be diluted below detection in bulk tissue^10,33^. Second, MDD is etiologically and pathologically heterogeneous^34^, and a mixed cohort of MDD cases, including those who died by suicide or not, with varying medication histories and illness durations, may dilute a consistent disease signal, given that MDD patients who died by suicide versus those who died from other causes show divergent, even opposing, transcriptomic profiles in prefrontal cortex^35,36^. Third, post-mortem brain studies are inherently limited by tissue quality, agonal state, pH, and PMI variability, which add noise^37^. Fourth, the absence of global GlycoRNA pathway difference suggests that the relevant pathology is not a global up- or down-regulation of glycosylation machinery per se, but rather an alteration in the coordination between this machinery and microglial state. This distinction, between absolute level and inter-pathway coupling, is precisely why interaction-based approaches (GEE, slope contrasts) were more informative than simple group comparisons^38,39^.

### 4.5 Methodological considerations: GEE as an exploratory framework

Traditional per-cell-type linear regression tests each GlycoRNA-cell-type relationship independently, without accounting for the fact that the four cell-type scores are measured from the same individual and are therefore statistically non-independent. GEE with subject-level clustering handles this repeated-measures structure explicitly^17^, enabling a single joint model that estimates: (i) the baseline GlycoRNA slope for the reference cell type (microglia) in controls; (ii) cell-type-specific deviations from that reference slope; (iii) diagnostic-group deviations; and (iv) the three-way interaction of all three factors. This approach yielded a clear, statistically significant GlycoRNA main effect on microglia (FDR = 0.006) that would have been obscured in a simple all-samples Pearson correlation (r = 0.282, P = 0.052, marginal), consistent with the well-documented gain in statistical power afforded by jointly modeling repeated within-subject measurements rather than analyzing them independently^40^. The GEE framework also allowed us to compute delta-method confidence intervals for predicted simple slopes and to apply sensitivity analyses with transformed outcomes, strengthening confidence in the robustness of the GlycoRNA-microglial association.

We acknowledge, however, that ssGSEA-based cell-type inference is an imperfect proxy for actual cell-type abundance or activation state. Marker gene sets, while curated from well-established CNS transcriptomic resources, may not fully capture microglial diversity (e.g., surveillant vs. activated vs. dystrophic states), and the GlycoRNA gene set used here represents a focused subset of glycosylation enzymes rather than a comprehensive GlycoRNA repertoire as defined by recent glycomics studies.

### 4.6 Limitations

Several limitations should be noted. First, the sample size (n = 48; 26 MDD, 22 controls) is moderate, and the power to detect three-way interactions, typically requiring hundreds of observations, is limited. The ∼61% slope attenuation observed here, while directionally consistent, should therefore be considered preliminary. Second, this is a secondary analysis of a single cohort requiring independent replication. Third, bulk transcriptomic inference cannot definitively localize GlycoRNA expression to microglia; single-nucleus RNA-seq or spatial transcriptomics is needed, and ssGSEA-based cell-type inference may not capture microglial diversity (surveillant vs. activated vs. dystrophic states). Fourth, medication exposure, suicide status, and illness chronicity are potential confounders not formally modeled. Fifth, the GlycoRNA gene set represents a focused subset of glycosylation enzymes rather than the comprehensive glycan repertoire defined by recent glycomic approaches. Finally, the cross-sectional post-mortem design cannot establish causality; the decoupling observed may be a consequence rather than a cause of MDD pathology.

## 5. Conclusions and future directions

In summary, our findings reveal that in the healthy human prefrontal cortex, GlycoRNA pathway transcriptional signature is specifically and significantly coupled to microglial transcriptional signature, a neuro-immune glycosylation axis that appears numerically attenuated in MDD. While the three-way diagnostic interaction did not reach formal statistical significance in this moderate-sized cohort, the convergent directional pattern across ordinary regression, GEE, sensitivity analyses, and microglia-focused dummy models supports the hypothesis that GlycoRNA-microglial transcriptional coordination may be disrupted in MDD DLPFC. Future studies should: (i) validate this relationship in larger independent post-mortem MDD cohorts with sufficient power for three-way interaction testing; (ii) employ single-nucleus RNA-seq to localize GlycoRNA pathway expression specifically to microglia; (iii) integrate glycomic profiling (e.g., mass spectrometry of N-/O-glycans) to link transcriptional glycosyltransferase signatures to actual glycan structures on microglial surfaces; (iv) stratify MDD cases by suicide status, medication exposure, and illness chronicity to identify which clinical dimensions drive the decoupling; and (v) experimentally test whether modulating microglial glycosylation (e.g., via sialyltransferase or galactosyltransferase inhibition) alters microglial phagocytic or inflammatory phenotypes in preclinical MDD models. The present work provides an exploratory framework for investigating neuro-immune glycosylation as a novel molecular dimension of MDD pathophysiology.

## 6. Acknowledgements

Financial support and sponsorship Not applicable.

## Conflicts of interest

The authors have no relevant financial or non-financial interests to disclose. The authors have no conflicts of interest to declare that they are relevant to the content of this article. The authors have no financial or proprietary interests in any material discussed in this article. All authors certify that they have no affiliations with or involvement in any organization or entity with a financial or non-financial interest in the subject matter or materials discussed in this manuscript.

## Data Availability statement

Raw transcriptomic data supporting this work are publicly available in NCBI GEO under accession GSE102556 (https://www.ncbi.nlm.nih.gov/geo/query/acc.cgi?acc=GSE102556), originally published by Labonté et al. (2017)^2^. Custom R analysis scripts generated for this study are available upon reasonable request. Processed intermediate score matrices are provided in supplementary material.

All data produced in the present study are available upon reasonable request to the authors.

All data produced in the present work are contained in the manuscript.

All data produced are available online.

## Ethical statement

This study represents secondary bioinformatic re analysis of publicly available de identified post mortem human brain transcriptomic data. Original human subject ethical approval and informed consent had been completed in the source study (GSE102556), originally published by Labonté et al. (2017)^2^; no additional ethical approval was required for present computational analyses.

## Author’s contributions

All authors have made substantial contributions to the work and meet the criteria for authorship as defined by the ICMJE (International Committee of Medical Journal Editors). All authors approved the final version to be published; agreed to be accountable for all aspects of the work to ensure that questions related to the accuracy or integrity of any part of the work are appropriately investigated and resolved. Their specific contributions are as follows:

Michael Maes:

❿ Supervision: Oversaw and took leadership responsibility for the research activity planning and execution.

❿ Conceptualization: Contributed to the ideas, formulation, and evolution of overarching research goals and aims.

❿ Writing – review & editing: Critically reviewed the manuscript and provided guidance on manuscript writing.

Rui Xu:

❿ Conceptualization: Contributed to the ideas, formulation, and evolution of overarching research goals and aims.

❿ Investigation: Performed literature search, review, and evaluation of relevant studies.

❿ Formal analysis: Conducted statistical analysis and interpretation of research results.

❿ Visualization: Prepared data visualizations.

❿ Writing – original draft: Prepared the initial draft of the manuscript.

❿ Writing – review & editing: Critically reviewed and revised the manuscript based on feedback.

Yingqian Zhang:

❿ Investigation: Performed literature search, review, and evaluation of relevant studies.

## Supporting information

Supplementary Figures 1_Microglia/Neuro/Oligodendrocyte/Astrocyte_boxplot

Supplementary Figures 2_GlycoRNA_Neuron/Astrocyte/Oligodendrocyte_cor

Supplementary Table 2_limma_DEG_GSE102556_final

Supplementary Table 3_group_comparison_descriptives

Supplementary Table 4_GlycoRNA_AllCellType_final_score

Supplementary Table 5_Table_Pearson_correlation_summary

Supplementary Table 6_two_way_interaction_lm

Supplementary Table 7_GEE_3way_coef_with_P_adjust

Supplementary Table 8_GEE_sensitivity_sqrt/log_transform

Supplementary Table 9_GEE_dummy_microglia_vs_others_2way/3way

## Notes

### Competing Interest Statement

The authors have declared no competing interest.

### Author Declarations

Raw transcriptomic data supporting this work are publicly available in NCBI GEO under accession GSE102556 (https://www.ncbi.nlm.nih.gov/geo/query/acc.cgi?acc=GSE102556), originally published by Labonte et al. (2017). Custom R analysis scripts generated for this study are available upon reasonable request. Processed intermediate score matrices are provided in supplementary material.

