## Supplementary figures and images for "Attenuated GlycoRNA-microglial transcriptional coupling in the dorsolateral prefrontal cortex of patients with major depressive disorder: an exploratory interaction-based in-silico analysis"

### Supplementary Figures 1_Microglia/Neuro/Oligodendrocyte/Astrocyte_boxplot

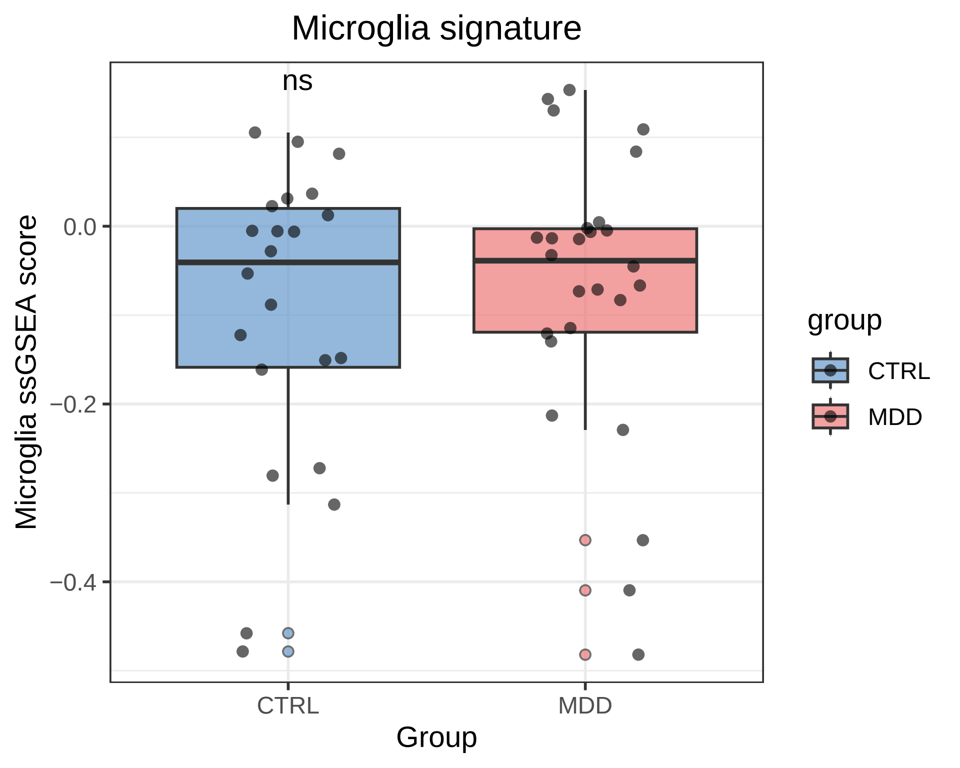

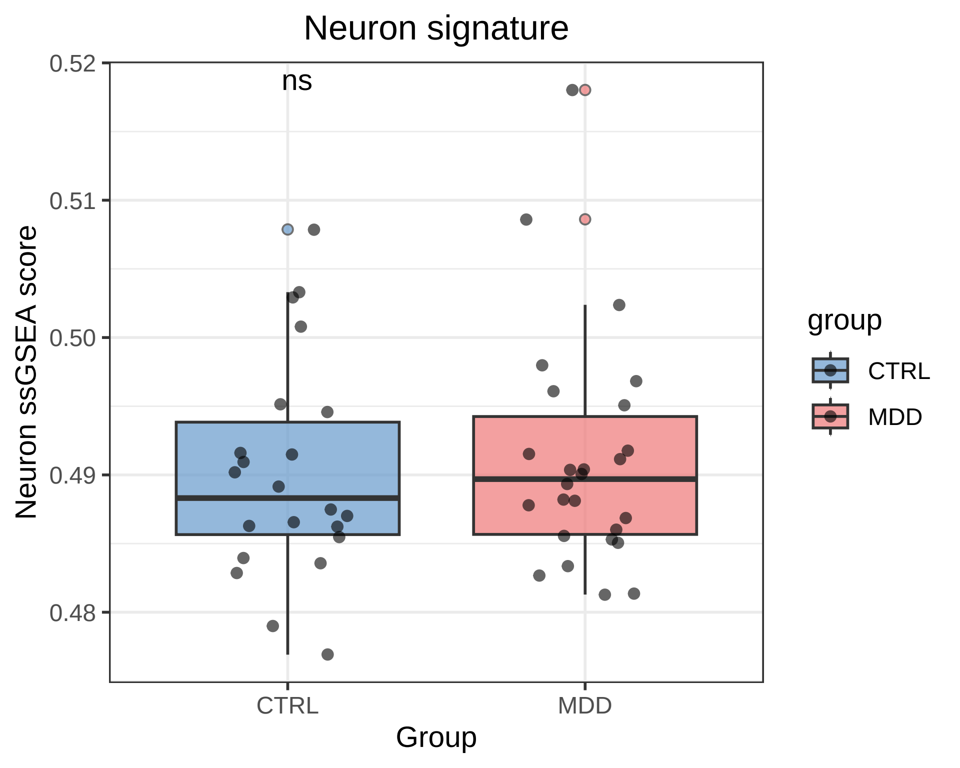

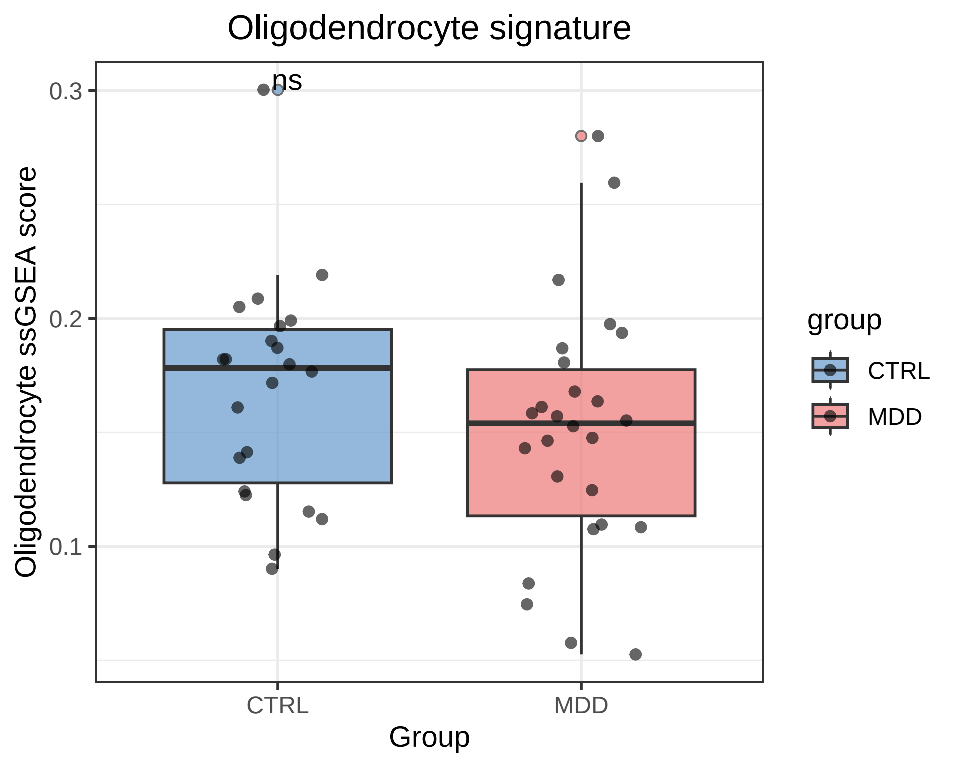

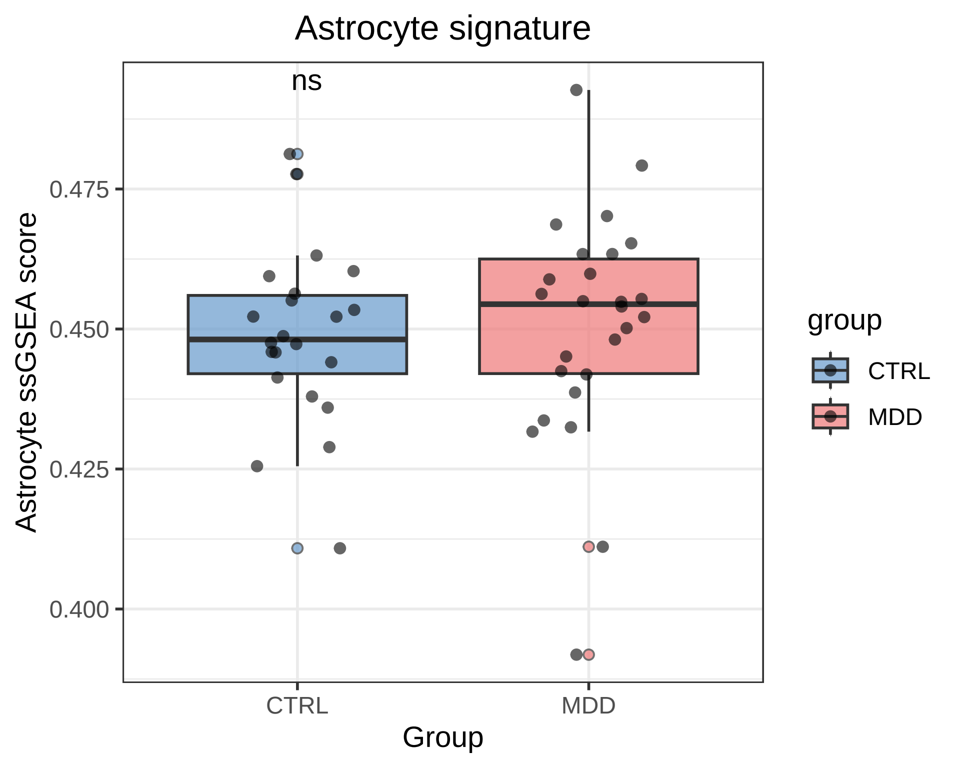

### Supplementary Figures 2_GlycoRNA_Neuron/Astrocyte/Oligodendrocyte_cor

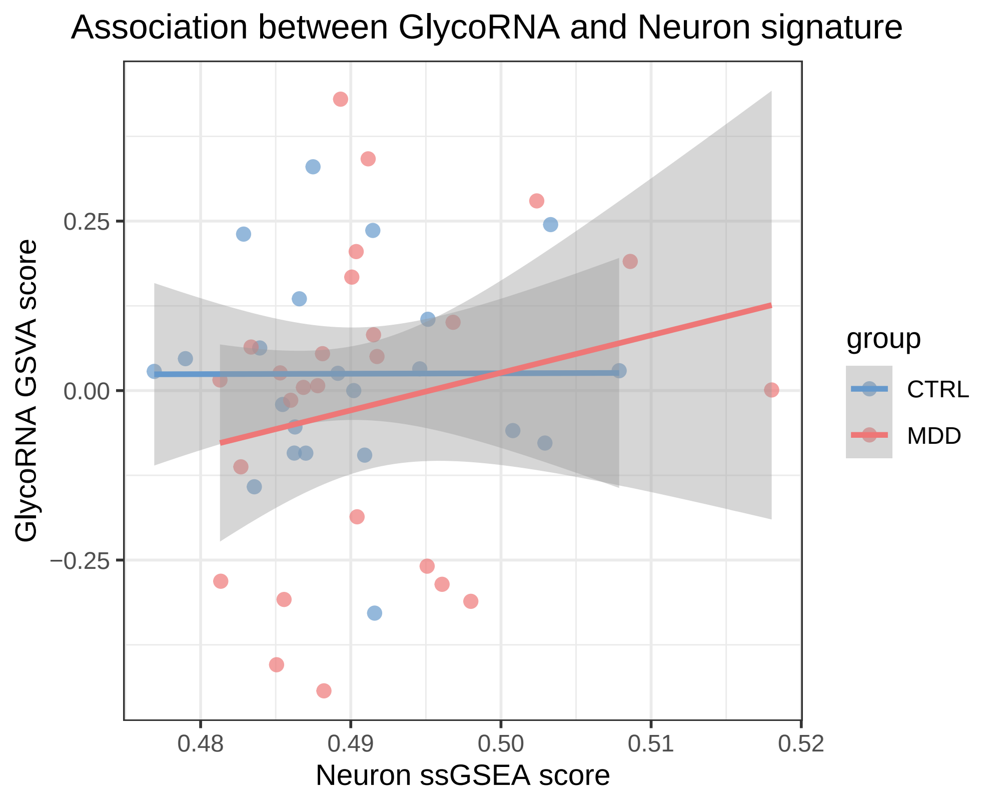

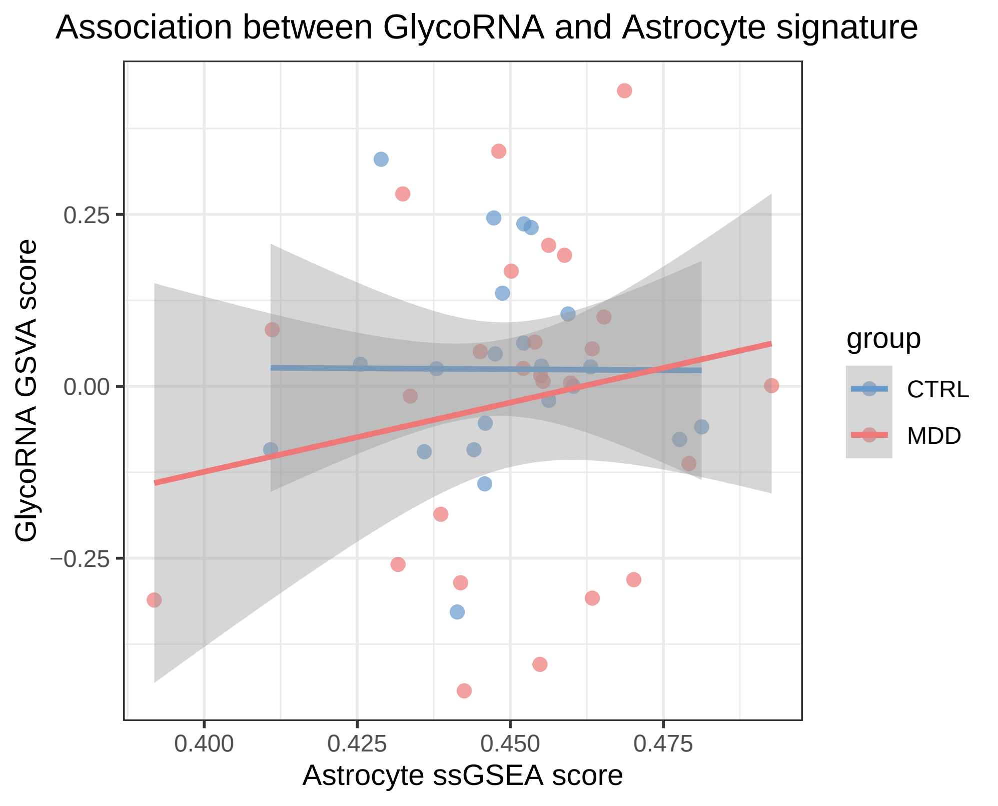

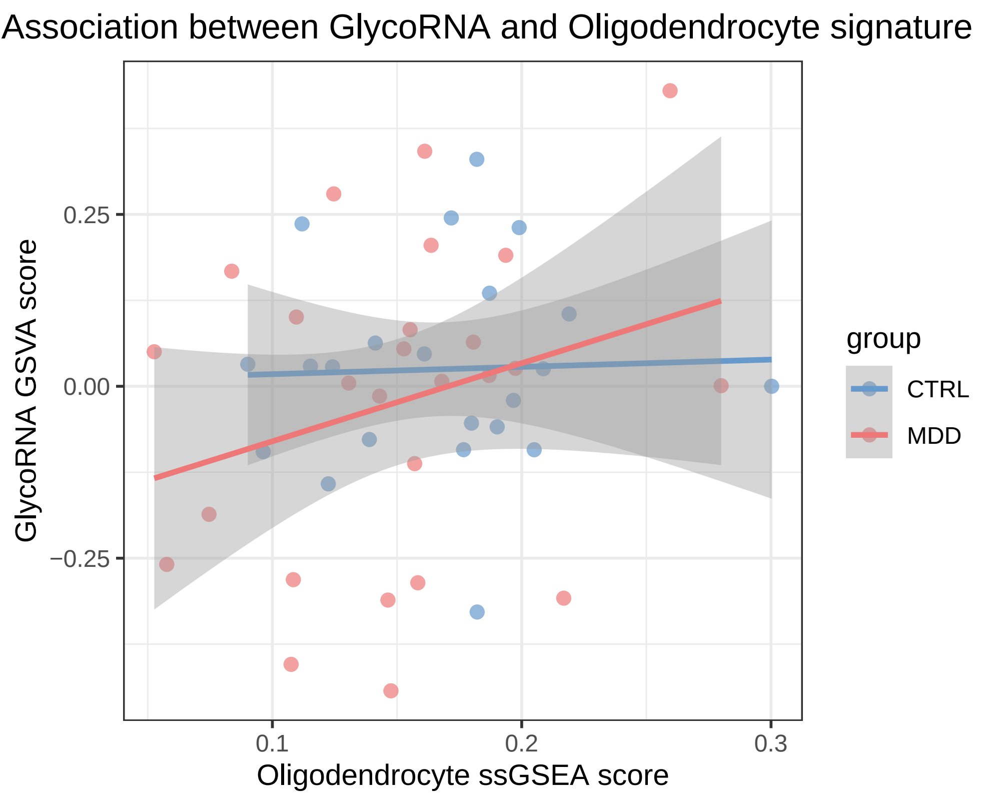
